# Somatic Mosaicism Patterns Define Clinical-Surgical Subtypes of Focal Cortical Dysplasia Through Cell-Type-Specific Expression

**DOI:** 10.1101/2025.10.06.25337172

**Authors:** Camila Araújo Bernardino Garcia, Muhammad Zubair, Xincen Xi, Ian Alfred Graham, Sang Hyun Lee, Sai Babu Patarlapalli, Marcelo Volpon Santos, Hélio Rubens Machado, Xiaoxu Yang

**Author notes:** These authors contributed equally to this work (Camila Araújo Bernardino Garcia, Muhammad Zubair). Correspondence (H.R.M.); (X.Y.).

## Abstract

Focal Cortical Dysplasia Type II (FCDII) is a subtype of cortical malfunction and is the primary cause of drug-resistant epilepsy in children. Although somatic mosaicism and clonal expansion of brain cells have been identified as crucial factors in FCD cases, the overall genetic landscape and clinical implications of FCDII remain largely unclear due to a significant gap in translating genetic data to inform surgical approaches and prognostic evaluations of individual cases. We carried out deep exome sequencing and deep amplicon validation of surgical biopsies and matched blood samples from 14 FCDII patients with confirmed neuropathology. We further performed multiscale pathogenic validations and took advantage of existing single-nucleus RNA sequencing and spatial maps from developing human cortices to explore the functionality of potential pathogenic somatic variants. We identified novel somatic variants in several functional categories, like neurotransmission (*TAAR2, GRM6, ZACN*), structural regulation (*TUBB2A, PLEC, COL18A1*), cellular maintenance (*IDO2, PARP4, P2RX5*), and RNA processing (*RBMX*), mapping the expression of these genes back to the developing human brain demonstrated significant enrichment in neuronal cell types, especially excitatory neurons, further confirming their contributions in early brain development and phenotypic functions in dysmorphic neurons. Combining these genetic findings with clinical phenotypes, we found brain-specific mosaic variants with very high mosaic fractions (fraction of mosaic cells, MF, up to 99.5% on *P2RX5*) associated with different clinical phenotypes. FCDIIB, a more severe subtype that contains balloon cells, had higher MFs (>40%) for variants within resectable cortical layers (excitatory neurons in Layers 5 and 6). This allows potentially targeted resection and achieves better clinical outcome (87.5 % with Engel score I). FCDIIA subtype, on the other hand, displayed lower MFs (<5%) with diffuse distribution, and required hemispherectomy, with poor surgical outcomes (Engel score II/III). Our results suggest MF thresholds are high-definition biomarkers of surgical outcome estimate, with MF > 40% predicting viable focal resection and MF < 5% indicating network dysfunction that necessitates broad-spectrum resection. Combining genetic mapping with cellular localization thus offers a coherent solution to precision surgery in FCDII, translating molecular diagnosis to clinical practice.

## Introduction

Focal Cortical Dysplasia Type II (FCDII) is a subtype of malformation of cortical development, which is one of the most frequent causes of drug-resistant epilepsy in children and young adults (Blümcke et al. 2011). Histopathological classification differentiates FCDIIA, which contains dysmorphic neurons but lacks balloon cells, and FCDIIB, which contains both dysmorphic neurons and balloon cells (Najm et al. 2018) as shown in subtypes such as tuberous sclerosis complex (TSC) (Grajkowska et al. 2010) and Hemimegaloencephaly (HME) (Arai et al. 1999). This histologic distinction correlates with the clinical presentation; however, the underlying biological processes are not fully elucidated. Recent evidence has established that somatic mutations during the process of cortical development are a vital pathogenic driver in FCDII (Baldassari et al. 2019; Chung et al. 2023). These genetic variants, which happen during the post-zygotic stage, cause a mosaic pattern of abnormal tissue formation in the developing cortex, interfering with critical cellular activities, such as neuronal migration, cortical layer formation, and cellular differentiation (Poduri et al. 2013; D’Gama et al. 2017). The sequence in which these mutational events occur during neurodevelopment appears to predetermine not only the histologic subtype, but also the clinical severity, with earlier occurring mutations leading to more widespread malformations and severe epilepsy phenotypes (Jamuar et al. 2014). The mTOR pathway genes (*MTOR, DEPDC5, TSC1, and TSC2*) are reported to be the major genetic contributors in the pathobiology of FCDs (De Angelis 1989; Baulac et al. 2015; Lim et al. 2015; Møller et al. 2016). Recently, additional genes along this line have been identified, such as *AKT3*, *PIK3CA*, and *RHEB*, which further expand the spectrum of mTOR-related disturbances in cortical development (Jansen et al. 2015; Chung et al. 2023).

In this study, we applied deep whole-exome sequencing and artificial intelligence-powered mosaic variant calling pipelines to identify likely pathogenic mosaic variants for FCDII in a clinical cohort beyond the above pathway. The variants were functionally mapped to the single-cell transcriptome and spatial layout of the developing human cortex (Qian et al. 2025). Our analysis demonstrates that the mutational landscape of FCDII is broader than these established pathways and includes new targets impacting a variety of biological processes. We found novel missense variants in *P2RX5,* indicating an epileptogenic role of purinergic signaling, and a variant of *COL18A1*, which may be related to extracellular matrix defects underlying the developing dysplasia. Similarly, other gene mutations were identified in our FCDII cohort, in genes related to cytoskeletal organization (*TUBB2A, PLEC*), cell signaling (*GRM6, TAAR2*), and DNA repair mechanisms (*PARP4 c.3116T>C, p.Ile1039Thr, DDX11: c.1221C>A, p.Ser407Arg*), further complementing the genetic basis of FCDII. The Mosaic fraction (MF%, fractions of cells carrying mosaic mutations) of these somatic variants emerges as a critical indicator of clinical phenotype: high MF variants (>40%) correlate with early infantile onset and focal, surgically resectable malformations characteristic of FCDIIB, whereas low MF versions (<5%) associate with later onset and diffuse network dysfunction typical of FCDIIA. Thus, the Mosaic fraction (MF%) threshold effect can be considered a biological factor to explore the conflicting surgical results between the two subtypes, where FCDIIB shows an 87.5% Engel Class I medical outcome, as opposed to 0% observed in FCDIIA.

This research provides a unified pathogenetic model of FCDII based on an integrated clinical, pathological, genetic, and spatial transcriptional model of disease pathology that links both known and novel determinants of cellular phenotype and the surgical resectability of this disease. Our study suggests that precision medicine applications in epilepsy surgery could be based on the complete characterization of genetic profiles, which can be used to select the best candidates to have targeted resections and help develop new treatment models of this drug-resistant condition.

## Results

### Divergent clinical profiles and surgical prognosis in FCDII subtypes and FCDIIB show earlier seizure onset but better surgical outcomes than FCDIIA

Clinical analysis of 14 patients who had surgical treatment of Focal Cortical Dysplasia Type II (FCDII) (Najm et al. 2018), and confirmed by histopathologic examination as either FCDIIA (n=6) or FCDIIB (6 FCDIIB, 1 with HME, and 1 with TSC), is distinct **(Figure 1A)**. Histopathological examination of resected brain tissue confirmed the diagnosis of FCDII in all 14 patients presented. As shown in **Figure 1B**, the microscopic analysis revealed the classic hallmarks of FCDII, including cortical dyslamination, and the presence of balloon cells and dysmorphic neurons. These findings provide the essential pathological basis for the subsequent genetic analysis conducted on this patient cohort. The selected cohort was male-enriched (10:4, M: F, p=0.1796, not significant by an exact binomial test), and the median age at surgery was 10.5 years **(Table 1)**. A key finding is that the age of onset of seizures varied significantly between subtypes. FCDIIB had a much earlier median onset of 5 months, many in infancy, compared to FCDIIA at a median of 5.8 years. This result was statistically confirmed by the given data (p=0.03504, single-tailed Mann-Whitney U test) **(Figure 1C)**. Interestingly, preoperative seizure frequency was extremely inconsistent, with the FCDIIB group having a larger median monthly seizure frequency (112.5 vs. 46.5), but not significantly so. Most patients (11/14, 78.6%) showed an associated neurological or developmental disability (ADNPM). There was a highly consistent and localizing pattern of pre-operative diagnostic information **(Table 1)**.

**Figure 1:**
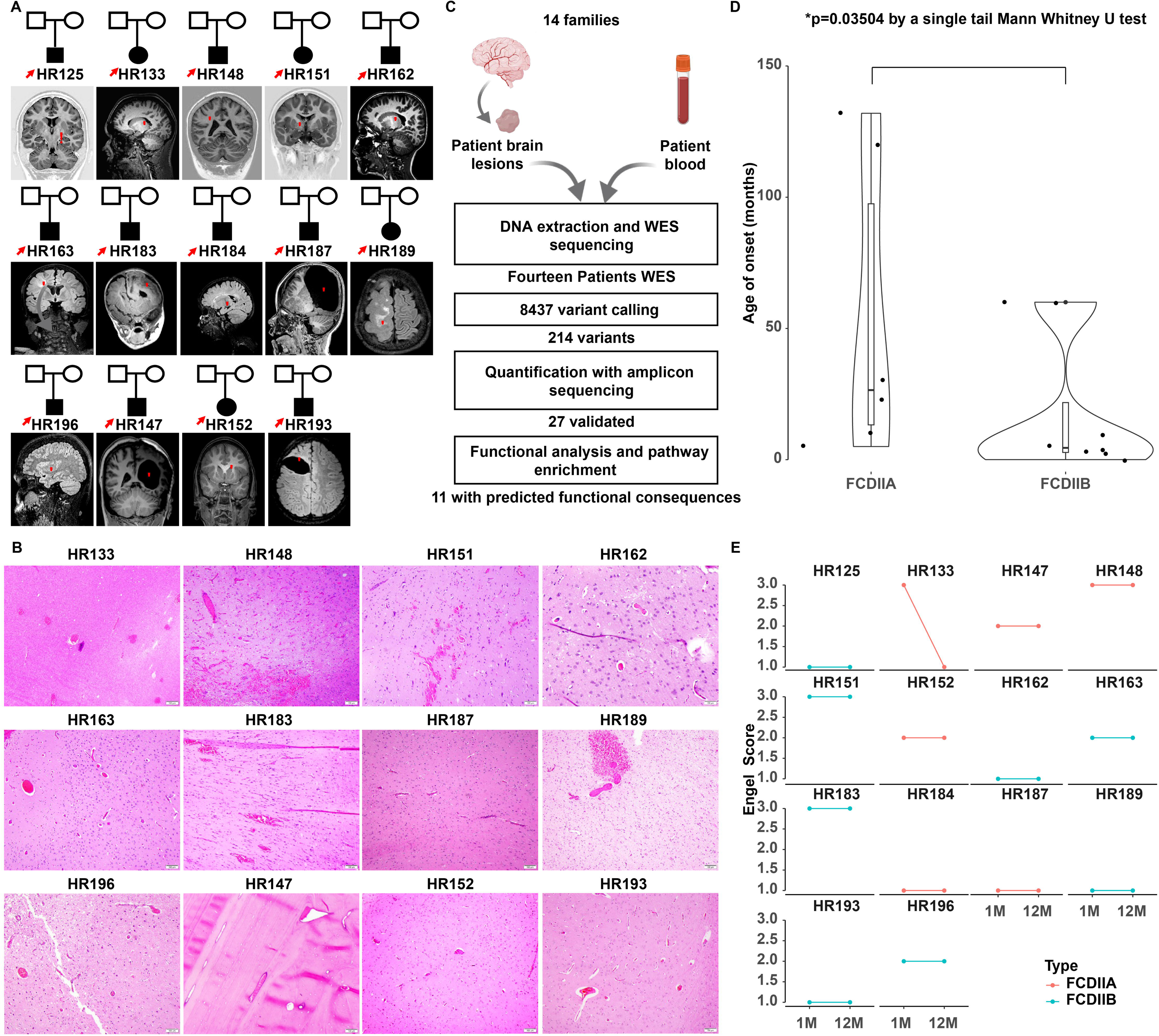
Cohort description, variant detection pipeline, and clinical overview. **(A)** Cohort characteristics and neuroimaging results: Fourteen patients diagnosed with focal cortical dysplasia type II (FCDII) were recruited in this study. Pedigree symbols: square (male), circle (female), dark symbols represent FCDII affected patients. Red arrows indicate patients included in whole-exome sequencing. The typical lesion patterns of the brain on MRI (indicated by red stars) are demonstrated as cortical thickness, loss of white-gray junction sharp identification, and transmantle sign. **(B)** Representative histopathological images of brain lesions from 14 patients with FCDII: Histopathological analysis of resected brain tissue from patients in the cohort confirms the diagnosis of Focal Cortical Dysplasia type II, revealing characteristic cellular and architectural abnormalities. Scale bars represent specific magnifications: 100 µm **(C)** Patient recruitment, experimental and computational pipeline, as well as orthogonal validation for potential pathogenic mosaic variants: Schematic of a multi-tiered bioinformatics workflow which includes sequencing data quality control (FastQC, MultiQC), alignment to the reference genome (BWA-MEM), removal of duplicates (Picard), paired-mode variant calling (Mutect2), and variant annotation (SnpEff, VEP). Filtering criteria included depth of coverage (≥30×), population frequency (<0.1% in gnomAD), and computational pathogenicity predictions. **(D)** Age of disease onset difference between FCDII subtypes: an earlier age of onset for epilepsy indicates a more severe phenotype (Berg et al. 2012). FCDIIB patients showed significantly earlier age of onset compared with FCDIIA patients (p=0.03504, single-tailed Mann-Whitney U test). **(E)** Assessment of Engel scores by type (FCDIIA and IIB) and time point. dotplots indicate the distribution of scores at two time points: one-month baseline (1M) and 12-month follow-up (12M).

**Table 1:** Clinical, radiological, and surgical Outcome characteristics of the FCDII cohort.

| Patient ID | Gender | Histopathology | Surgery Type | Lobe | Onset | Pre-op Sz Freq (/mo) | ADNPM | Hemiparesis | Pre-op MRI Key Finding | Engel 1M | Engel 1Y |
| --- | --- | --- | --- | --- | --- | --- | --- | --- | --- | --- | --- |
| HR125 | M | FCD IIB | Lobectomy | Temporal | <=5Y | 135 | Y | N | Cortical blurring (R temp/par) | 1 | 1 |
| HR133 | F | FCD IIA | Hemispherotomy | Temporal | 5-10Y | 150 | N | N | Cortical thickening (temporal) | 3 | 1 |
| HR147 | M | FCD IIA | Hemispherotomy | Hemispheric | <=5Y | 90 | Y | Y (HH) | WM volume loss | 2 | 2 |
| HR148 | M | FCD IIA | Hemispherotomy | Fronto-Parietal | 11-15Y | 60 | N | Y | FLAIR hyperintensity (L frontal) | 3 | 3 |
| HR151 | F | FCD IIB | Hemisph.-Lobect. | Temporal | <=5Y | 180 | Y | Y | WM asymmetry, cortical indistinction | 3 | 3 |
| HR152 | F | FCD IIA | Hemispherotomy | Parietal | <=5Y | >104 | Y | Y | WM-GM blurring (frontal) | 2 | 2 |
| HR162 | M | FCD IIB (Tuberous Scl.) | Hemispherotomy | Temporal | <=5Y | 90 | N | N | Cortical tubers (temporal) | 1 | 1 |
| HR163 | M | FCD IIB | Lobectomy | Temporal | <=5Y | Uncont | Y | N | Polymicrogyria | 2 | 2 |
| HR183 | M | FCD IIB | Lesionectomy | Temporal | <=5Y | 14 | Y | Y | Hemimegalencephaly | 3 | 3 |
| HR184 | F | FCD IIA | Lobectomy | Temporal | <=5Y | 150 | Y | N | Hippocampal asymmetry | 1 | 1 |
| HR187 | M | FCD IIA | Lesionectomy | Temporal | <=5Y | 210 | Y | N | Ventricular dilation, dysplasia | 1 | 1 |
| HR189 | M | FCD IIB (HME) | Hemispherotomy | Frontal | <=5Y | 100 | Yes | No | Hemispheric hypertrophy | 1 | 1 |
| HR193 | M | FCD IIB | Hemispherotomy | Frontal | <=5Y | 750 | Y | N | Cortical thickening (frontal) | 1 | 1 |
| HR196 | M | FCD IIB | Lesionectomy | Frontal | <=5Y | Uncont. | Y (ADHD) | N | Focal cortical thickening | 2 | 2 |

- Histopathology: FCD (Focal Cortical Dysplasia), HME (Hemimegalencephaly), Tuberous Scl. (Tuberous Sclerosis Complex), Pre-op Sz Freq: Monthly seizure frequency pre-operation; "Uncont." denotes uncontrollable/day-to-day variability, ADNPM (Associated Disability/Neurological/Psychological Morbidity): Y (Yes), N (No). Specifics (e.g., ADHD, Hemianopia), Hemiparesis: Y (Yes), N (No). HH = Homonymous Hemianopia, Engel Scale: 1 (Seizure-free), 2 (Rare seizures), 3 (Worthwhile improvement), Surgery Type: Hemisph.-Lobect. = Hemispherotomy-Lobectomy, Pre-op MRI Key Finding, R/L = Right/Left; Temp = Temporal; Par = Parietal; WM = White Matter; GM = Grey Matter, ADHD: Attention-Deficit/Hyperactivity Disorder (neurodevelopmental condition, often comorbid with epilepsy ; HH: Hypothalamic Hamartoma (brain lesion associated with epilepsy and behavioral disturbances)

Abnormalities of the EEG were reported to exist in all 14 of the patients who had definite reports, and the results were able to correlate precisely with the localization of the epileptogenic area in all cases (Jamuar et al. 2014). EEG was characterized by disorganized baseline activity, regional slowness, and epileptic paroxysms (e.g., sharp waves, polyspikes), which were most common in the affected hemisphere. However, patient HR133 (FCDIIA) had an irritative state of the right temporal lobe in the area of the abnormal seizure onset zone, and patient HR148 (FCDIIA) had paroxysmal activity of epileptiform nature in the left parasagittal region **(Table 1)**. In 93% (13/14) of cases, pre-operative MRI was also diagnostic, with only one FCDIIB patient (HR196) having a not-so-severely abnormal pre-operative scan. The most typical examples of MRI findings were thickening of the cortical area (e.g., HR193, HR163), blurring of the grey and white matter borders (e.g., HR151, HR152), and a hyperintense signal of the subcortical white matter on T2/FLAIR (e.g., HR148, HR162). The transmantle sign, which is a hallmark characteristic of FCDII, was expressly present in patient HR133. The most frequent procedure was hemispherotomy (7/14), which was used in more extensive lesions **(Table 1)**.

A dramatic difference was illustrated in the surgical outcome at both one month and one year at follow up as shown in the longitudinal outcome figure. Eighty-seven percent of the patients in whom an Engel Class I outcome (seizure-free) was reached had the FCDIIB subtype. The results also showed that these excellent results in FCDIIB patients were already observed at the 1-month postoperative and were consistent at 12-month follow-up. In contrast, none of the FCDIIA patients were seizure-free, with distributions in Engel Class II (50%, rare seizures) and Class III (50%, worthwhile improvement) **(Figure 1D)**. This stratification is visualized clearly in the graphical data, where FCDIIB patients are consistently clustering in lower Engel scores at both timepoints (1.0-1.5), whereas FCDIIA patients show more variable and higher scores (2.0-3.0). This pictorial representation, coupled with the statistical significance of the difference in the outcomes between subtypes, converges to augment the statistical significance of a difference between subtypes **(Figure 1C)**. In short, FCDIIB occurs at an earlier age and is found with impressively good surgical results that are determined early and are stable over time, whereas FCDIIA occurs later, with much less favorable post-operative activity results. The electro-clinical-radiological correlations were robust with EEG and MRI effectively localizing the lesion for the surgical planning of the patient, and the longitudinal outcome provided clear evidence of the durable therapeutic response of completely resected FCDIIB cases.

### Somatic mutation burden correlates with severity and surgical prognosis in Focal Cortical Dysplasia Type II

Next-generation deep exome sequencing of FCDII patient brain tissue and control samples **(Figure 1B)** demonstrated a novel somatic mutational landscape, which establishes a molecular explanation of its clinical heterogeneity, and 27 potential somatic mutations **(Figure 2A-C)** were observed in 14 patients, and the mutational charge was also greater in FCDIIB, averaging 2.6 compared to 1.8 mutations per patient in FCDIIA. Some FCDIIB cases presented with multiple co-existing mutations; notably, patient HR193 had five mutations (*SMPDL3B*, *DISP1*, *OR10A2*, *IDO2*, *PLEC*). FCDIIA cases generally reported one or two variants in comparison **(Table S1)**. The identified somatic variants genes coincided with important functional pathways; cytoskeletal/structural integrity, where a truncating loss of function variant in *TUBB2A* (a neuronic beta-tubulin, HR152-FCDIIA) and a loss of function variant in *PLEC* (a cytoskeletal linker, HR193-FCDIIB) were identified, cell signaling/receptors (*GIPR*, *GRM6, TAAR2*, *P2RX5*) and DNA repair/regulatory elements (*TP53BP1*, *DDX11*, *PARP4*) **(Figure 2B&C)**. This genetic data directly describes the clinical phenotypes as outlined earlier.

**Figure 2:**
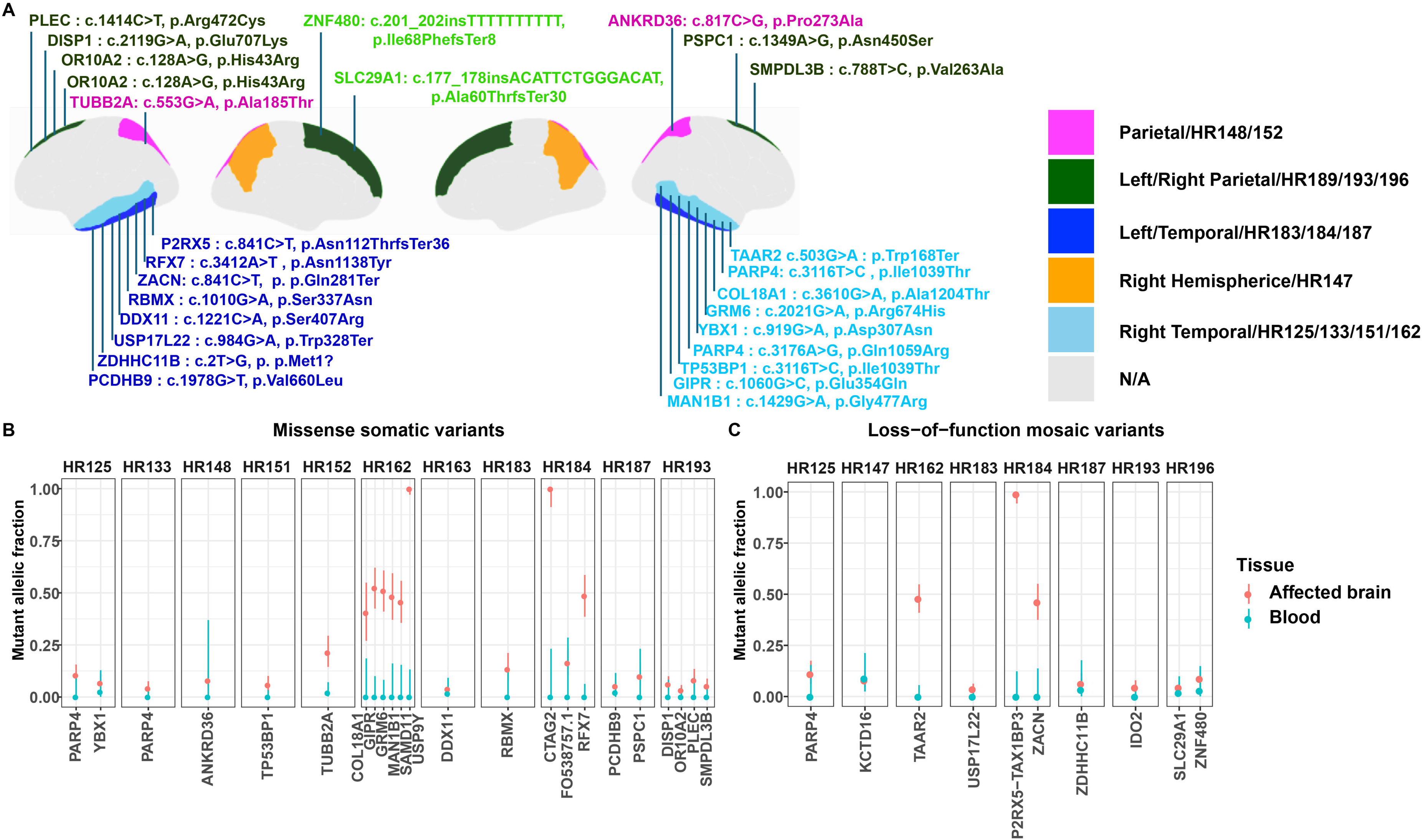
Novel missense and loss-of-function somatic variants landscape associated with FCDII, detected and quantified by WES. **(A)** Anatomical distribution of somatic variants: The variants visualized in color are tracked to their affected brain regions with patients’ lesions. **(B)** Dot plot of missense mosaic variants and their Mutant Allelic Fractions with 95% binomial confidence intervals, arranged by donor ID. Tissue types are color-coded **(C)**. Frameshift mosaic variations with mutant allelic fraction values and their 95% binomial confidence intervals: Color codes are the same as (B).

The increased burden of mutations in FCDIIB, which interfere with pathways that regulate cortical development as well as the cortical cytoskeleton organization, is perfectly in line with its much earlier seizure onset (median 5 months) and with the severe malformations (e.g., cortical thickening, transmantle sign). The less mutated profile in FCDIIA and targeting functions specialized at a later developmental age point to a later presentation (median 5.8 years) and significantly less favorable surgical outcomes (Engel Class II/III), indicating a possibly more diffuse/elusive epileptogenic zone **(Figure 1C & Table 1)**. The difference in Engel Class I outcome, found only in FCDIIB patients, may thus be explained by the relative selectiveness of a highly mutagenic lesion irreversibly causing early developmental changes in a localized area that is fully resectable, as opposed to an undifferentiated pattern of developmental disturbance in FCDIIA **(Figure S1A&B)**.

Moreover, the somatic nature of the variants was substantially confirmed by low Mutant Allelic Fractions (MAF), which is a typical factor of post-zygotic mutations **(Figure 2B&C)**. In summary, molecular-clinical integration reveals that FCDIIB is associated with an increased somatic mutation burden in various developmental pathways, correlating with the severe early-onset disease with favorable early resection outcomes. FCDIIA exhibits the simpler mutational makeup that is related to later onset and worse surgical outcomes. The relevant genes implicate the dysregulation of neuronal migration, cytoskeletal organization, and cell signaling as the central disease processes in FCDII.

### Validated pathogenicity of somatic variants, including early clonal drivers, underpins the FCDII clinical spectrum

The pathogenic potential of identified somatic variants was also confirmed comprehensively through multifaceted analysis. In silico characterization with CADD(Kircher et al. 2014) Phred scores **(Figure 3A&B, Figure S1C-F)** indicated high deleterious frank: *TAAR2* (p.Trp168Ter, CADD=38) and *ZACN* (p.Gln281Ter, CADD=38) protein-truncating variants. The functional effect was further estimated using the GPN-MSA(Frazer et al. 2021; Benegas et al. 2025) score, where the measure of an evolutionary constraint evaluates the rarity of a specific amino acid change across a multiple sequence alignment. A highly negative GPN-MSA score suggests a high likelihood of functional and good probability of evolutionary conservation. Some of the variants indicated intensely negative scores, thus providing an affirmation of their destructive effects. These were *GRM6* (p.Arg674His, GPN-MSA=-10.34), *TAAR2* (p.Trp168Ter, GPN-MSA=-10.32), *COL18A1* (p.Ala1204Thr, GPN-MSA=-9.37), *RFX7* (p.Asn1138Tyr, GPN-MSA=-9.11), *TUBB2A* (p.Ala185Thr, GPN-MSA The combination of high CADD and strongly negative GPN-MSA scores strongly indicates the pathogenic nature of these variants **(Figure S1C&D)**.

**Figure 3:**
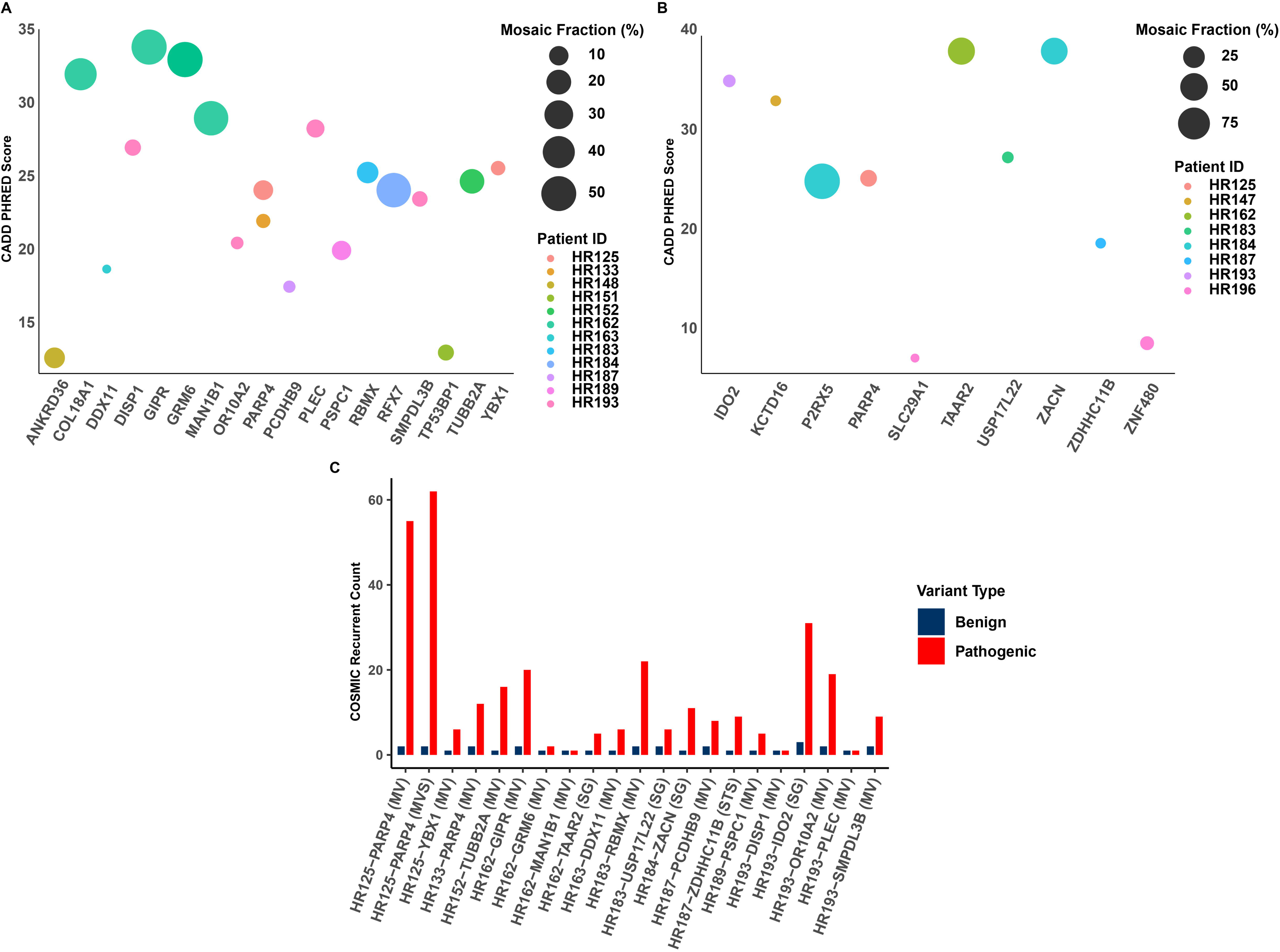
Predicted high-impact variants exhibit higher mosaic fractions (MF%) and greater recurrence in COSMIC. **(A)** Mosaic fraction correlation: Relationship between CADD scores and mosaic fraction for missense somatic variants, demonstrating higher pathogenicity scores in high MF variants. **(B)** Parallel analysis for loss-of-function somatic variants showing similar trends. **(C)** COSMIC recurrence analysis: Bar plot showing recurrence counts of identified somatic variants in cancer databases compared to benign polymorphisms in the same genes.

Prior occurrence in Somatic Cancer Databases Cross-mapping against the Catalogue of Somatic Mutations in Cancer (COSMIC)(Tate 2010) revealed that many of these variants were recurrent somatic cancer mutations, highlighting their significance as cancer drivers. Notable ones are a *PARP4* variant (p.Ile1039Thr) that has been observed 62 times by COSMIC, an *IDO2* stop-gained variant (p.Tyr346Ter) with a 31 recurrence count, and an *RBMX* missense change (p.Ser337Asn) that has been recorded 22 times **(Figure 3C)**. The repetitive nature of cancer gives strong external evidence of its functional importance and oncogenic potential, which is very relevant to the dysregulated cortical development.

The somatic origin and clonal distribution of a variant were conclusively determined by estimating the Mosaic Fraction (MF% %)(Mo and Walsh 2025; Zubair and Yang 2025), which varied broadly (2.2-99%). This spectrum is characteristic of post-zygotic mosaicism. A higher frequencies of MF% values were detected in a group of variants, including the frameshift in *P2RX5* (p.Asn112ThrfsTer36, MAF=99%), missense in GRM6 (MAF=50.9%), and *RFX7* (MAF=48.5%) **(Figure 3A&B)**. These high percentages indicate that these mutations were very early in embryonic development, so they would have resulted in numerous clones and are most likely the cause of the focal cortical malformation. In contrast, smaller MF% values (e.g., *DDX11* at 2.2% imply subsequent mutational events. This investigation of pathogenicity as multiple parameters has an inherent connection with the established clinical hierarchy of FCDII. The FCDIIB subtype was not only defined by a more severe mutational burden, but also a distribution of more strongly pathogenic variants: a higher CADD score, stronger evolutionary selection against it, and a greater level of recurrence in cancer databases. Moreover, a few FCDIIB cases (e.g., HR162, HR184) showed extremely high mosaic frequencies, suggesting early developmental age and extensive clonal expansion, which is consistent with the extremely widespread malformations observed on MRI (e.g., transmantle sign, hemimegaloencephaly) **(Table 1& S1)** and corresponds to their good surgical outcomes upon focal driver resection. In contrast, FCDIIA variants were also found to be pathogenic, but the lower overall mutational burden and the lack of very high MF% likely caused a less complete developmental disruption, later seizure age, and less resectable epileptogenic zone, possibly explaining the less favorable surgical outcomes**(Figure 1C&D, Figure S1A&B)**. Accordingly, the CADD, GPN-MSA, COSMIC, and MF% data could be combined to develop an explanatory model of somatic variants’ molecular pathogenicity in the context of FCDII with direct clinical and phenotypic classification.

### Cellular and spatial architecture of somatic variants defines a neurodevelopmental hierarchy underlying surgical outcomes in FCDII

Single-cell transcriptomics and spatial analyses enable the exploration of these mosaic variants in normal developing human brain to understand the mechanisms causing FCDII (Baldassari et al. 2025). Combining snRNA-seq data (human fetal brain (Qian et al. 2025), GW20) and spatial transcriptomics (Visium and MERFISH) showed that gene variants do not occur randomly in expression, but rather are concentrated in specific, essential cell populations (Nowakowski et al. 2017). The UMAP visualization of the integrated dataset outlined the primary cerebral cell types, including excitatory neurons **(Table 2)** (ExN), inhibitory neurons (InN), glia (astrocytes, oligodendrocytes), and radial glial (RG) progenitors **(Figure 4A&B, Figure S2)**.

**Figure 4:**
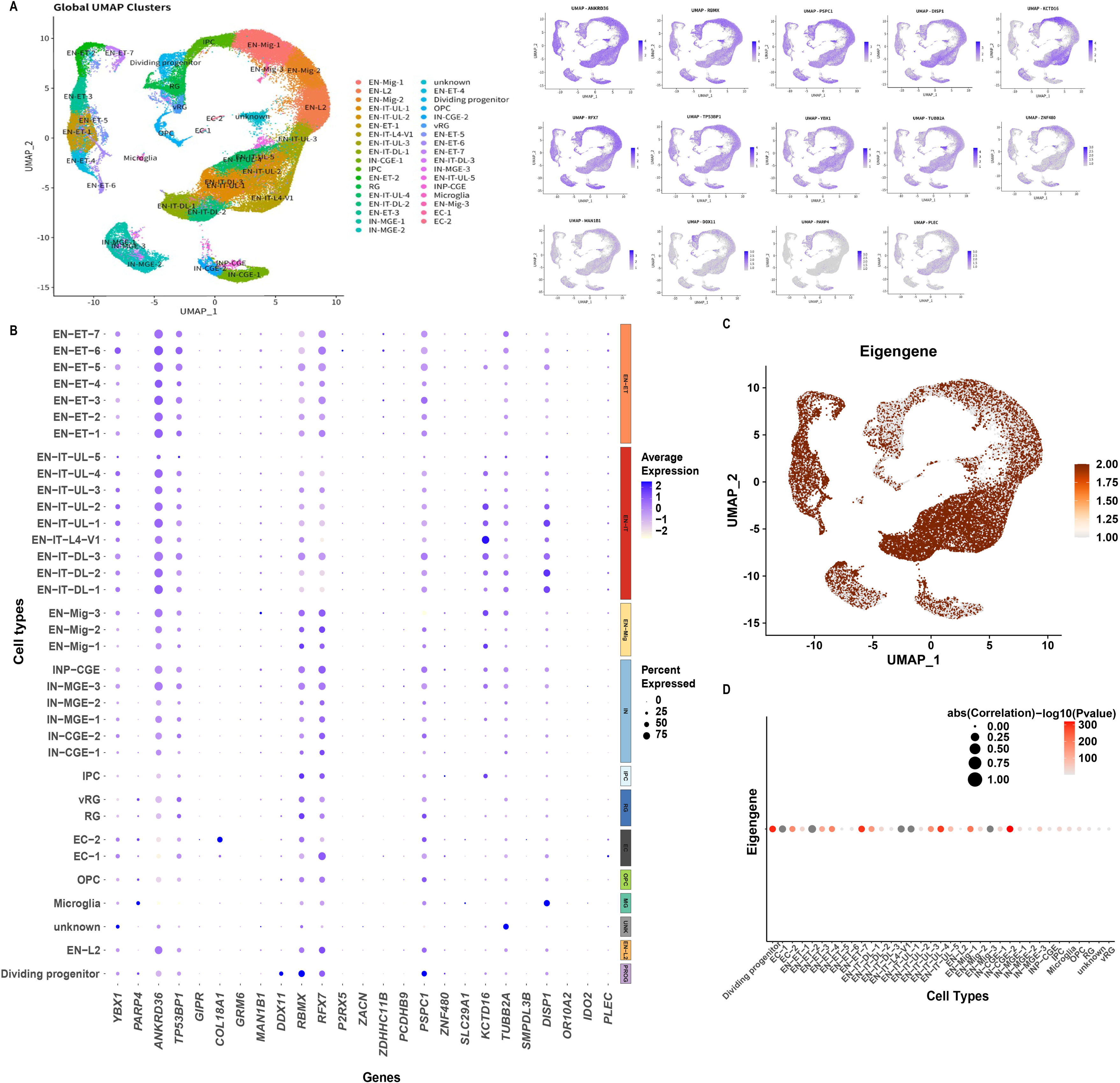
Mapping genes with somatic mutations in FCDII in single-cell transcriptomics and spatial architecture of the developing human brain. **(A)** Integrated single-nucleus RNA sequencing (snRNA-seq) and spatial transcriptomics reveal cell-type-specific enrichment patterns of potential somatic variant-bearing genes. UMAP visualization of major cerebral cell types from integrated fetal (GW20) and adult cortical datasets, showing enrichment of FCDII-associated genes in specific populations: excitatory neurons (ExN-L5/6), inhibitory interneurons (InN-CGE), radial glial progenitors (RG), and oligodendrocyte precursor cells (OPCs). **(B)** Guided by spatial informatics, transcriptomic profiles of variants detected from the genes mutated in FCD2 patients were mapped to developmental human cortexes to explore the spatial and cell-type enrichment. Each row corresponds to a cell type cluster, and each column corresponds to a candidate gene. The size of the dot reflects the percent of the specific gene in the specific cell type. The intensity of the dot indicates the average expression level. Color blocks on the right indicate cell type groups **(C)** Eigengene analysis of the overall enrichment of the FCD2 expressed gene mapped back to the developmental human brain. The UMAP dimensions are the same as Figure 4A. **(D)** Correlation of the eigengene analysis in each individual cell type. The size of the dots indicates the correlation coefficient. The intensity of the dots indicates the negative log-transformed p-value of the correlation. Both Figure 4 C and D showed that the genes found mutated in FCDII brains are significantly enriched in neuronal cell types in the developing human brain.

**Table 2:** Potential candidate somatic variants in the FCDII cohort.

| Gene | Variant | MF (%) | CADD | GPN-MSA | COSMIC Recurrence | Epilepsy Enrichment | Primary Cell Type Enrichment | FCD Subtype | Resectability | Engel Outcome Link |
| --- | --- | --- | --- | --- | --- | --- | --- | --- | --- | --- |
| TAAR2 | p.Trp168Ter | 47.9 | 38 | -10.32 | 5 | Yes | ExN-L5/6, InN | FCDIIB | High | Engel I |
| ZACN | p.Gln281Ter | 46.2 | 38 | -7.86 | 11 | Yes | ExN-L5/6, Progenitors | FCDIIB | High | Engel I |
| GRM6 | p.Arg674His | 50.9 | 33 | -10.34 | 2 | Yes | ExN-L5/6, OPCs | FCDIIB | High | Engel I |
| COL18A1 | p.Ala1204Thr | 40.4 | 32 | -9.37 | - | Yes | ExN-L5/6, RG | FCDIIB | High | Engel I |
| P2RX5 | p.Asn112ThrfsTer36 | 99 | 24.9 | - | - | Yes | Diffuse glial/progenitor | FCDIIA | Low | Engel II/III |
| TUBB2A | p.Ala185Thr | 19.6 | 24.7 | -9.2 | 16 | Yes | ExN-L5/6, RG | FCDIIA | Moderate | Engel II |
| RBMX | p.Ser337Asn | 13.4 | 25.3 | -6.71 | 22 | Yes | InN-CGE, OPCs | FCDIIB | Moderate | Engel I |
| IDO2 | p.Tyr346Ter | 4.5 | 35 | -6.99 | 31 | Yes | Progenitors, glia | FCDIIB | Low | Engel I |
| PARP4 | p.Ile1039Thr | 11 | 25.2 | -5.36 | 62 | Yes | InN-CGE, OPCs | FCDIIB | Moderate | Engel I |
| PLEC | p.Arg472Cys | 8.1 | 28.3 | -8.09 | 1 | Yes | ExN-L5/6, RG | FCDIIB | High | Engel I |

Analysis of variant gene expression revealed strongly specific and distinctive patterns of enrichment across these clusters. The key genes, which include *TUBB2A*, *GRM6*, and *PLEC*, reflected an enriched expression in deep-layer excitatory neurons (ExN-L5/6), which is a critical component of cortical output and circuit formation. Additional genes were also enriched in specific subtypes of interneurons derived from caudal ganglionic eminence (InN-CGE) and oligodendrocyte precursor cells (OPCs), including *RBMX* and *PARP4* **(Figure 4A&B, Figure S2)**. The expression of various variants in RG progenitors and upper-layer excitatory neurons (ExN-L2/4) highlighted the developmental timing and cellular origins of the malformations (Mayer et al. 2018; Breuss et al. 2022). To evaluate the overall expression pattern of the candidates in developing human cortex, we constructed an eigengene analysis described previously(Chung et al. 2023). An eigengene cluster representing the expression pattern of the candidate gene as a group is mapped to the developing human brain **(Figure 4C)**, and the relative correlation of the eigengene cluster to each cell type was calculated **(Figure 4D)**. The high correlation (r = [e.g., 0.82], p < [e.g., 1e-200]) demonstrates that the overall expression pattern of this eigengene module is strongly associated with neuronal clusters, especially the excitatory neurons.

To further explore the enrichment of the above candidate genes against epileptogenic situations, orthogornal phenotyping was performed against the CellxGene database (Program et al. 2025). Particularly, *PSPC1*, *ZNF480*, *PLEC*, *KCTD16*, *TUBB2A,* and *DISP1* genes were highly overexpressed in temporal lobe epilepsy **(Figure 4B**, **Table 2)**. The gene RBMX, which we identified as mutated in our cohort, provided particularly high enrichment (expression values 1.5-4.0) to the epileptic tissue we used, and it encodes an RNA-binding protein involved in RNA processing and neuronal activity.

Likewise, *MAN1B1* and *ZDHHC11B* exhibited increased expression (3.7-4.0), which is in line with their being involved in seizure pathogenesis **(Figure S2A)**. Spatial transcriptomics has been used to localize this cellular enrichment to specific anatomical contexts. The variants with high mutant allelic fraction (MF% >40%), found in our foregoing analysis as early developmental drivers, in particular, were localized to specific, focal regions of cortical architecture. This anatomical focality, especially in clearly defined laminar anatomy like ExN-L5/6, offers a direct mechanistic rationale to their successful surgical resection and the consequent excellent Engel Class I outcomes in FCDIIB patients following anatomically targeted resection. In contrast, variants with lower MF% (<5%) and FCDIIA have more diffuse expression, enriched in glial and progenitor cells that are less anatomically constrained **(Figure S2B)**.

These network modulating variations with extensive cellular expressions show correlation with a less resectable nature and worse surgical outcomes that often require more extensive hemispherotomy than others to achieve Engel Class I outcomes. Spatial representation in a single cell combines ideally with the foregoing genomic and clinical discoveries that underpin the therapeutic value of focality in FCDIIB, in which high MF% variants are strictly resectable, entailing targeted resections, whereas the progenitor/glial diffuseness of the variants in FCDIIA reflects the underlying imperviousness to therapy, necessitating hemispherotomy to achieve seizure-free status**(Figure S2)**. Further validation provided by the epilepsy gene expression databases supports further biological validity of our candidate genes in the pathogenesis of seizures **(Figure 5A)**. The MF% threshold (>40%) could therefore serve as an essential biomarker in the surgical plan, and could be a multi-scale**(Figure 5B&C)** pathophysiological model of somatic mutation, cellular phenotype, and surgical resectability.

**Figure 5:**
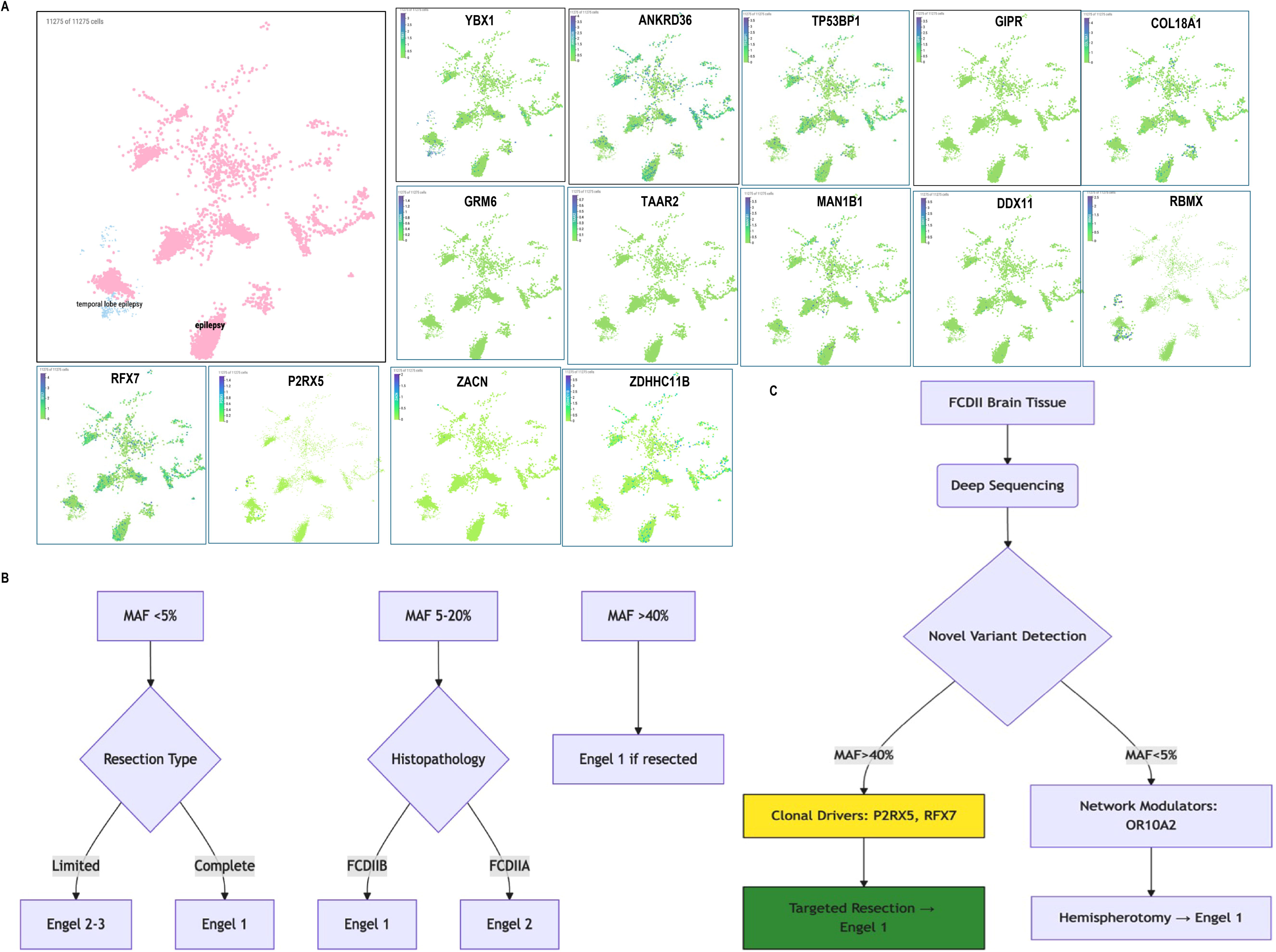
Mosaic fraction and cellular specificity as biomarkers for surgical outcome prediction in FCDII patients. **(A)** The integrated potential identified the FCDII gene with spatial architecture with molecular data, emphasizing colocalization of pathogenic gene expression to elucidate region-specific mechanisms in epileptogenesis. **(B)** Correlation between mosaic fraction thresholds and surgical outcomes: The figure demonstrates that variants with MF >40% (predominantly FCDIIB) are localized to surgically resectable layers and show excellent Engel Class I outcomes (87.5%), while variants with MF <5% (FCDIIA) display diffuse expression patterns and poorer surgical outcomes (0% Engel Class I). **(C)** Clonal dominance predictive value supporting their utility as biomarkers for surgical stratification.

## Discussion

FCDII reflects a subset of cortical malformations arising due to somatic mutations during brain development (Steriade et al. 2021; Najm et al. 2022), with the ILAE classification distinguishing FCDIIA (dysmorphic neurons) and FCDIIB (balloon cells and dysmorphic neurons)(Garcia et al. 2025). This study presents a multiscale analysis of Focal Cortical Dysplasia Type II (FCDII), combining neuropathological-clinical, genomic, pathogenicity, and spatial transcriptomic data to create a unified model of its pathogenesis and treatment response. As our results suggest, these subtypes constitute distinct clinical genetic entities with radically different patterns of developmental origin, mutational architecture, and epileptogenicity circuits that directly determine the effects of surgical resectability and clinical outcomes.

The dramatic clinical difference between the subtypes is deep-rooted in their somatic mutational landscapes. FCDIIB has been characterized by an increased burden of developmentally relevant pathogenic mutations (mean 2.6 vs. 1.8 per patient) in the pathways of cytoskeletal organization (*PLEC*, *TUBB2A*), cell signaling (*GRM6*, *TAAR2*), and DNA repair (*PARP4*, *DDX11*). These preliminary post-zygotic mutations (MAF >40% in driver genes such as *GRM6* and *TAAR2*) lead to general clonal expansion and severe focal malformations as seen on MRI (e.g., transmantle sign, cortical thickening), and subsequent infantile-onset seizures. FCDIIA, on the other hand, has a less populated assortment of mutations (MAF <5% in genes such as *ANKRD36*), higher-frequency later-acting variants, with more particularized functions, which allow lateralized insult onset and create diffuse, network-wide malfunctioning (Table 3).

**Table 3:**
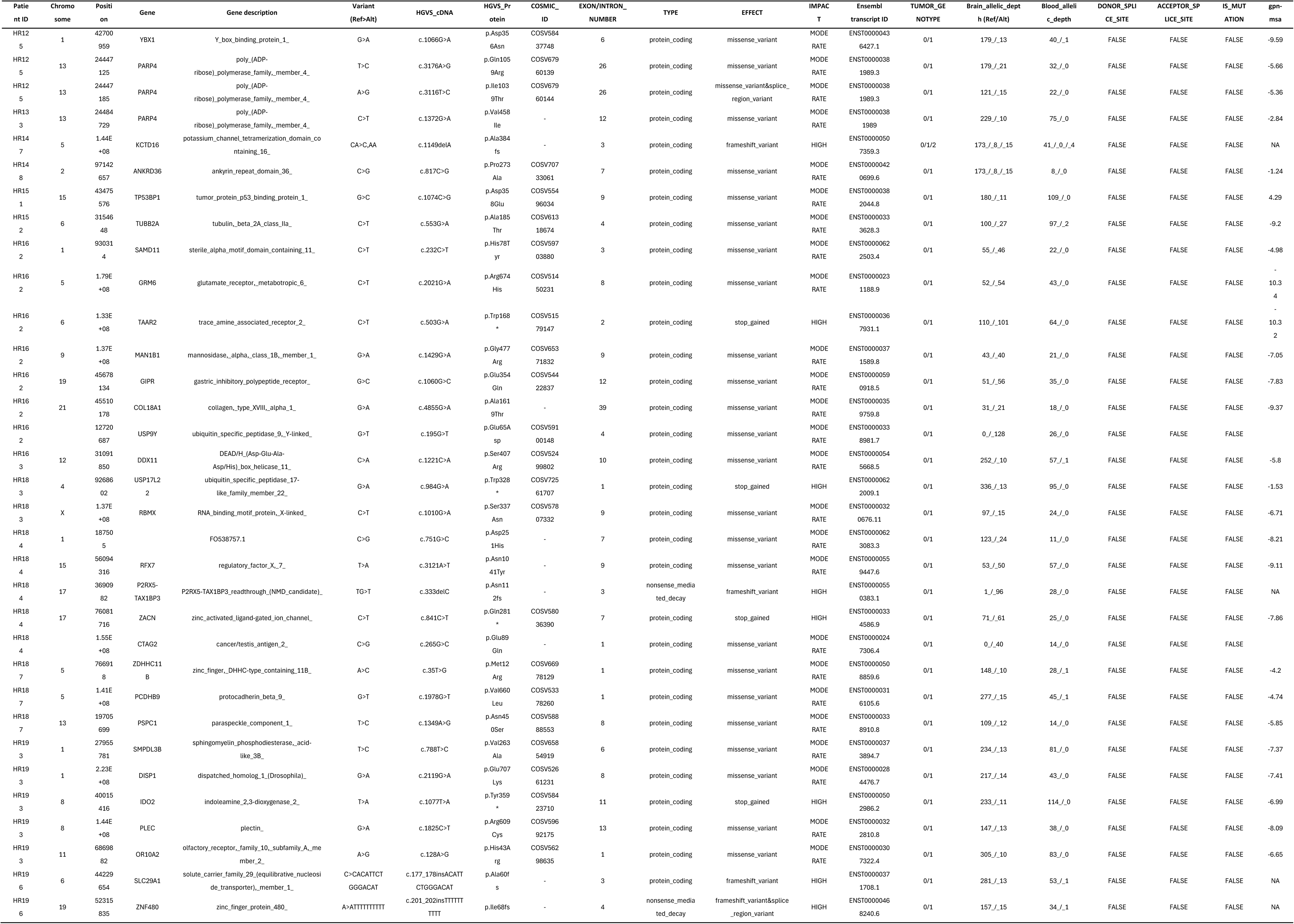
Candidate mosaic variants and their functional predictions.

More importantly, multi-parametric pathogenicity verification demonstrated the biological feasibility of these variants. Their pathogenicity was supported by a high CADD score (*TAAR2* p.Trp168Ter=38), extreme evolutionary pressure (GPN-MSA score -9 for GRM6, TUBB2A), and cancer recurrence (*PARP4* p.Ile1039Thr in 62 COSMIC entries), which contributed to their potential in cortical abnormality disruption. The somatic nature was undisputably established by allelic fractions congruent with mosaicism (1-99%). Critically, multi-parametric pathogenicity validation made it biologically plausible that these variations represented an active pathogenicity via high CADD scores, substantive evolutionary constraint (GPN-MSA <-9), and cancer database recurrence. The mosaic fraction became a critical operative biomarker, and thresholds (>40%, 5-40%, <5%) dictated surgical treatment and outcomes. High MF variants (>40%) were associated with predisposition to focal expression in resectable cortical layers and a favorable prognosis in terms of surgical success, whereas low MF variants (<5%) were characterized by focal expression, associated with more extensive resection. Further Spatial transcriptomics brought the last connection between genotype and surgical outcome: anatomical focality. The FCDIIB (*GRM6*, *PLEC*) variants reached high-MAF and localized to discrete and resectable stratums (ExN-L5/6), which facilitated a resection with Engel I outcomes. In contrast, low-MAF FCDIIA variants were diffusely expressed in progenitors/glia, forming poorly defined epileptogenic connectivity in need of hemispherotomy to achieve seizure freedom. The very strong correlation between eigengene significance and neuronal cell types **(Figure 4D)** indicates that the detected genes in FCD2 patient brains are key members in neuronal functions and significantly correlate with the clinical trait. This positions them as high-priority candidates for further functional validation. The cell type enrichment of the identified genes aligns with the known pathophysiology of FCD, providing biological plausibility for our findings. External validation using CellxGene verified the overexpression of these genes (*RBMX*, *TUBB2A*) in epilepsy and supports their role in causing seizures **(Figure 5A)**. Our results support a genetically driven surgical approach in which mosaic fraction mapping guides precision surgery. Preoperative genetic screening, especially high-MF (>40%) variants, has the potential to maximize patient selection in targeted resection **(Figure 5B&C)**. The novel mechanisms identified raise the possibility of targeted interventions against purinergic signaling (*P2RX5*) and extracellular matrix defect (*COL18A1*).

## Limitations

The current limitation of small sample size on identified somatic variants needs further study and functional validation on model organisms (e.g., lineage tracing organoids/CRISPR-Cas9) to foster a causal mechanism that could strengthen the clinical framework.

## Conclusion

Our study suggests a modified model of FCDII pathogenesis, whereas FCDIIB is a product of early somatic mutations with high cellular burden, forming focal and well-defined malformations, which can be mostly removed by surgical excision, and is a good indication of excellent seizure control. Comparison-wise, FCDIIA is seen with a later mutation that is less prevalent in cells affecting the network dysfunction, and it is less surgically removable. This leads to FCDIIB providing better response to surgical procedures, and FCDIIA exhibiting poor response to surgical procedures, given its infiltrative nature. Moreover, beyond the mTOR pathway, we find novel variants in neurotransmission (*TAAR2: c.503G>A: p.Trp168Ter* (Revel et al. 2011)*, GRM6: c.2021G>A, p.Arg674His* (Elia et al. 2012)*, ZACN: c.841C>T, p. p.Gln281Ter* (Peralta and Huidobro-Toro 2016), structural regulation (*TUBB2A: c.553G>A, p.Ala185Thr* (Jaglin et al. 2009)*, PLEC: c.1414C>T, p.Arg472Cys* (Vahidnezhad et al. 2022)*, COL18A1: c.3610G>A, p.Ala1204Thr* (Garn et al. 2025)), cellular maintenance (*IDO2: c.1038T>A, p.Ala185Thr* (Kaswan et al. 2025)*, PARP4 c.3116T>C, p.Ile1039Thr* (Schreiber et al. 2006)*, P2RX5: c.841C>T, p.Asn112ThrfsTer36* (Chessell et al. 2005), and RNA processing (*RBM: c.1010G>A, p.Ser337Asn* (Liu et al. 2022). The combination of molecular characterizations of novel somatic variants with the clinical and imaging data will have the potential to improve patient outcomes by providing better guidance of the surgery and ultimately targeted appropriate medical therapy.

## Material and Method

### Clinical investigation of FCDII participants

Fourteen patients with Focal Cortical Dysplasia Type II (FCDII) **(Figure 1A)** were recruited in this study in the University Hospital of Ribeirao Preto Medical School (HCFMRP-USP). The complete presurgical assessment was undertaken in all the participants, including magnetic resonance imaging (MRI) and video-electroencephalographic (EEG) monitoring to precisely identify the epileptogenic zone **(Table 1)**. Surgery was advised when a localized, regional, or hemispheric epileptogenic zone was identified. Venous blood samples and brain tissue specimens pairs were collected during surgery to facilitate genetic mapping of mosaic variants.

### Ethical compliance

This research was approved by the Research Ethics Committee of HCFMRP-USP (Protocol No. 6979/2015). All participants had written informed consent forms signed by them or their authorized representatives before their inclusion in the study (Garcia et al. 2025). Raw genomic sequence data are controlled following IRB regulations.

### Tissue preparation and H&E staining

Operational specimens were snap-frozen in liquid nitrogen and stored at −80 °C. For analysis, samples were thawed at room temperature, and a tissue block (approximately 2 mm × 5 mm × 5 mm) was dissected orthogonal to the surface(Poduri et al. 2012). The blocks were fixed in 10% buffered formalin for 24 hours, dehydrated through a graded ethanol series, and embedded in paraffin. Sections were cut at a thickness of 3 μm and mounted on glass slides. For hematoxylin and eosin (H&E) staining, slides were air-dried at 60 °C for 20 minutes(Garcia et al. 2020). The paraffin was then removed with xylene, and the sections were rehydrated through a descending ethanol gradient. Nuclei were stained with hematoxylin, followed by differentiation in 0.3% hydrochloric acid alcohol and bluing in 0.2% ammonium hydroxide. Cytoplasm was counterstained with eosin. Finally, sections were dehydrated, cleared in xylene, and images were captured on an ultra-high resolution Olympus SZX16 stereomicroscope and processed with the CellSens software.

### Whole exome and amplicon sequencing and bioinformatics analyses

Exome sequencing was undertaken to detect disease-associated mosaic variants according to the scheme explained in **Figure 1B**. Raw sequencing data were evaluated with FastQC (Andrews 2010) and MultiQC(Ewels et al. 2016) with trimadaptor duplicate in TrimGalore (https://www.bioinformatics.babraham.ac.uk/projects/trim_galore/). Aligned reads were removed, then mapped against the human reference genome (GRCh38/GRCh37) using Burrows-Wheeler(Li and Durbin 2009) Aligner (BWA-MEM). After alignment, the processed files were marked as duplicates using Picard Tools (https://broadinstitute.github.io/picard/), and the coverage metrics were calculated. Variant calling was performed using Mutect2 and subsequent annotation through SnpEff (Cingolani et al. 2012), Universal Mutation Database (Béroud et al. 2000), and custom Perl scripts. These were filtered down (initially) to those with a minimum coverage of >=30 in normal tissue in the annotated variants. To prioritize pathogenic variants, we mimicked the potential consequences of the variants, including missense variants, splice donor/acceptor variants, stop gained/lost variants, start lost variants, TF binding site variants, 5 prime UTR premature start codon gain variants, and stop retained variants, using the reference genome mentioned above(McKenna et al. 2010). Mosaic variants were further validated and quantified using amplicon resequencing as previously described (Garcia et al. 2020), and the mutant allelic fractions, along with the 95% binomial confidence intervals, are provided in the supplementary tables.

### Somatic variant identification and mosaic fraction quantification

Functional annotation of variants was carried out with the help of VEP and ANNOVAR, which incorporated data from several publicly available databases (OMIM, ClinVar, HGMD, dbSNP, 1000 Genomes, ESP, ExAC, and gnomAD) (McKenna et al. 2010; Hunt et al. 2022). The given variant prioritization selection was performed using a multilevel filtering process that included: CADD scores (Rentzsch et al. 2019); population allele frequency; POLYPHEN and SIFT predictions; GPN-MSA deep-learning assessments (Benegas et al. 2025). The mosaic fraction was estimated as: MF = 2 VAF, where: Variant Allele Frequency (VAF) = (ALT reads in tumor)/(Total reads in tumor). Somatic origin was supported by the lack of alternative alleles on a matched blood sample (ALT = 0) (Mo and Walsh 2025) (Garcia et al. 2025).

### Single-nuclei RNA sequencing and spatial transcriptomics integration

Published single-nuclei RNA sequencing and spatial transcriptomic data from developing human cortex were retrieved in public repositories such as: SRA (PRJNA1231045) (Qian et al. 2025), dbGaP (phs000989.v3.p1) (Nowakowski et al. 2017), GEO (GSE162170; PMID: 34390642), and NeMO Archive (RRID: SCR_002001) The set of included data points used snRNA-seq profiles in fetal prefrontal (FB121_F1, GW20) and occipital cortices (FB080_O1/O2, GW20), along with Visium spatial profiles (FB080-O1 occipital cortex; FB121-F1 prefrontal cortex) and MERFISH (UMB1031-O1). The cell type-specific expression patterns were displayed using UMAP projections followed by manual curation of cell type annotations. Distributions of spatial gene expression were projected into anatomical contexts based on tissue slice images. Eigengene analysis was performed for the list of genes detected from the FCDII patients, as previously described(Chung et al. 2023), and mapped to the same dataset. The analysis code is available publicly at: https://github.com/XiaoxuYangLab/FCD2.

Besides, Spatial transcriptomic enrichment analysis for epilepsy(Program et al. 2025) and temporal lobe epilepsy was analyzed using CZ CELLxGENE (Program et al. 2025), a platform to conduct scalable discovery, analysis, and modeling of aggregated single-cell omics data.

## Data Availability

All data produced in the present study are available upon reasonable request to the authors

## Acknowledgements

The authors thank the patients and their families for participating in this study. The authors thank Dr. Kristen Kwan for providing the imaging instrument. This work was supported by PRONAS (25000.069257/2015-00) (To C.A.B.G. and H.R.M.) and NIH/NICHD R00HD111686 and NIH/NIMH R21MH134401 (To X. Y.). The support and resources from the Center for High Performance Computing at the University of Utah are gratefully acknowledged. The computational resources used were partially funded by the NIH Shared Instrumentation Grant 1S10OD021644-01A1.

## Author contributions

Conceptualization: C.A.B.G., M.Z., H.R.M., and X.Y.; methodology: C.A.B.G. I.A.G. and M.Z.; software: X.X., M.V.S., S.H.L., I.A.G., S.B.P., and X.Y.; validation: C.A.B.G. and M.V.S.; formal analysis: C.A.B.G. and M.Z.; data curation: C.A.B.G. I.A.G. and M.V.S.; writing—original draft: M.Z.; writing—review and editing: M.Z., X.Y., and C.A.B.G.; visualization: M.Z. X.X., and X.Y.; project administration: C.A.B.G., H.R.M., and X.Y.; funding acquisition: H.R.M., C.A.B.G., and X.Y. All authors have read and agreed to the published version of the manuscript.

## Code and data availabilities

The code used for plotting is available on https://github.com/XiaoxuYangLab/FCD2. All data produced in the present study are available upon reasonable request to the authors

## Conflict of interest

The authors declare no conflicts of interest related to this work.

---

**Figure S1:**
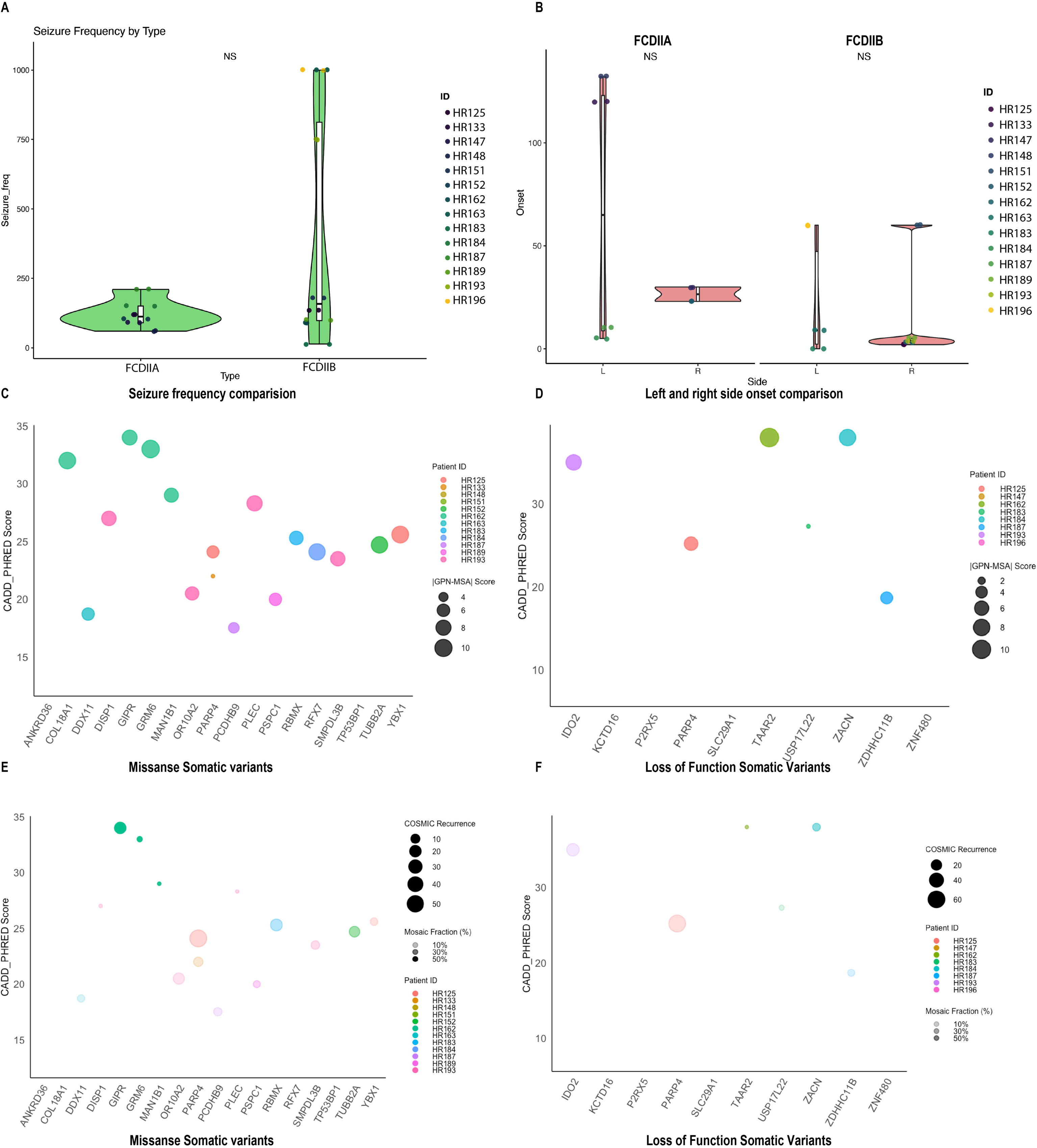
Correlation of clinical phenotype and features of the mosaic mutations detected from FCD patients. **(A&B)** The box and violin plots show the frequency of seizure onset (A) and landscape position on the brain region (B), R: Right, L: Left. The "NS" notations indicate that no statistically significant difference (p > 0.05) between the groups compared. **(C)** Missense somatic variant validation: Plot of CADD Phred scores (x-axis) versus GPN-MSA evolutionary constraint scores (y-axis) for missense variants. Circle size represents mosaic fraction, color indicates patient IDs; (CADD >20, GPN-MSA <-5). **(D)** Loss-of-function somatic variant validation: Similar analysis for protein-truncating variants showing stronger constraint signatures. **(E&F)** Functional analysis of CADD, MF%, and COSMIC scores of missense variants and loss-of-function variants.

**Figure S2:**
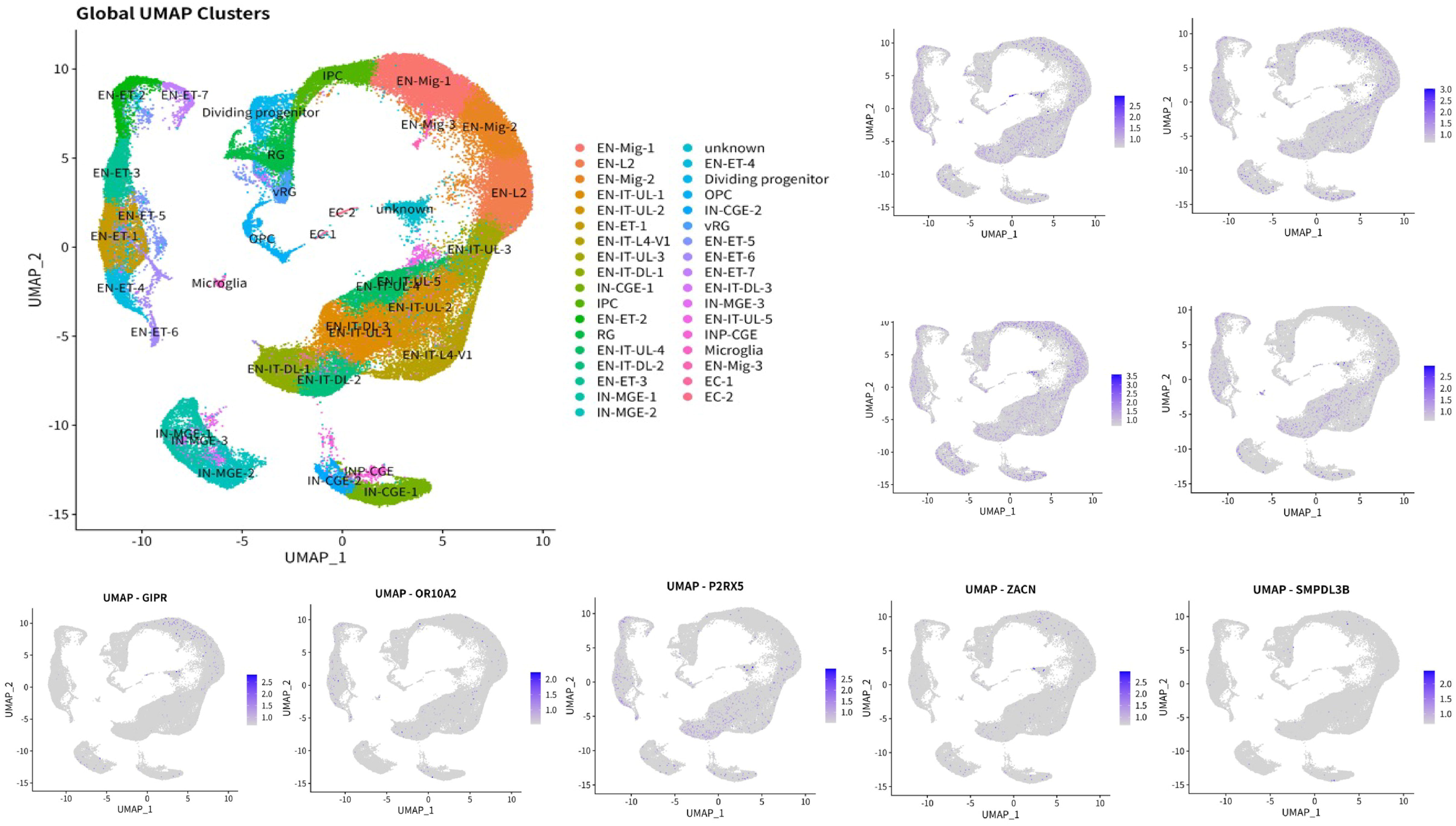
Cell type expression of genes with somatic variants detected from FCDII patient brains. High-resolution spatial mapping of remaining variants has lower expression within cortical microenvironments, showing preferential enrichment in specific cellular niches, including ventricular/subventricular zones, cortical plate, and developing white matter tracts. Left: global UMAP clusters with spatial information define the single-cell cell typing from different cortical layers in the developing human brain. Right: expression of mutated genes in the developing human cortex.

**Table S1:**
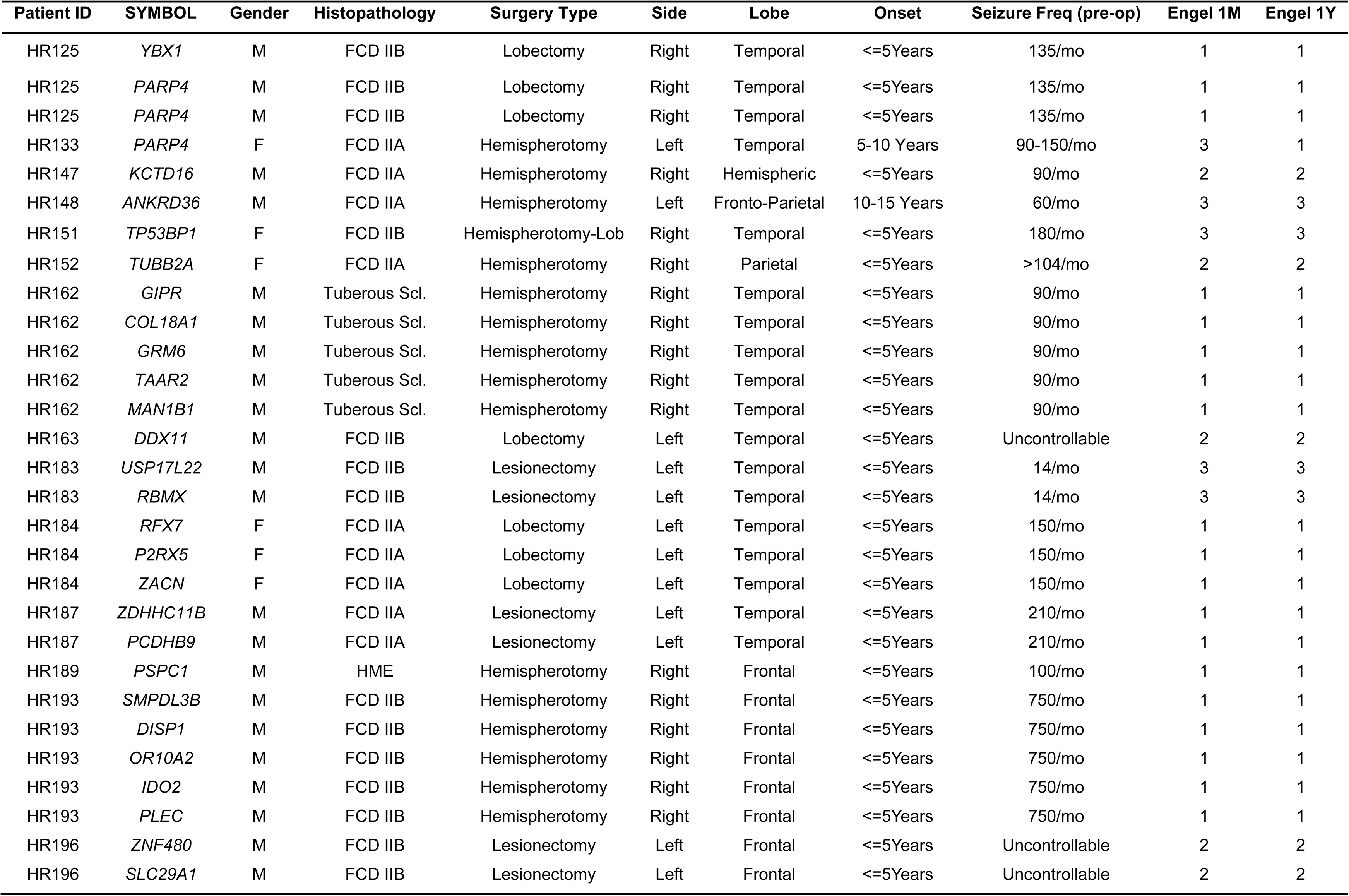
Clinical investigations of patients with identified somatic variants.

| Patient ID | SYMBOL | Gender | Histopathology | Surgery Type | Side | Lobe | Onset | Seizure Freq (pre-op) | Engel 1M | Engel 1Y |
| --- | --- | --- | --- | --- | --- | --- | --- | --- | --- | --- |
| HR125 | *YBX1* | M | FCD IIB | Lobectomy | Right | Temporal | <=5Years | 135/mo | 1 | 1 |
| HR125 | *PARP4* | M | FCD IIB | Lobectomy | Right | Temporal | <=5Years | 135/mo | 1 | 1 |
| HR125 | *PARP4* | M | FCD IIB | Lobectomy | Right | Temporal | <=5Years | 135/mo | 1 | 1 |
| HR133 | *PARP4* | F | FCD IIA | Hemispherotomy | Left | Temporal | 5-10 Years | 90-150/mo | 3 | 1 |
| HR147 | *KCTD16* | M | FCD IIA | Hemispherotomy | Right | Hemispheric | <=5Years | 90/mo | 2 | 2 |
| HR148 | *ANKRD36* | M | FCD IIA | Hemispherotomy | Left | Fronto-Parietal | 10-15 Years | 60/mo | 3 | 3 |
| HR151 | *TP53BP1* | F | FCD IIB | Hemispherotomy-Lob | Right | Temporal | <=5Years | 180/mo | 3 | 3 |
| HR152 | *TUBB2A* | F | FCD IIA | Hemispherotomy | Right | Parietal | <=5Years | >104/mo | 2 | 2 |
| HR162 | *GIPR* | M | Tuberous Scl. | Hemispherotomy | Right | Temporal | <=5Years | 90/mo | 1 | 1 |
| HR162 | *COL18A1* | M | Tuberous Scl. | Hemispherotomy | Right | Temporal | <=5Years | 90/mo | 1 | 1 |
| HR162 | *GRM6* | M | Tuberous Scl. | Hemispherotomy | Right | Temporal | <=5Years | 90/mo | 1 | 1 |
| HR162 | *TAAR2* | M | Tuberous Scl. | Hemispherotomy | Right | Temporal | <=5Years | 90/mo | 1 | 1 |
| HR162 | *MAN1B1* | M | Tuberous Scl. | Hemispherotomy | Right | Temporal | <=5Years | 90/mo | 1 | 1 |
| HR163 | *DDX11* | M | FCD IIB | Lobectomy | Left | Temporal | <=5Years | Uncontrollable | 2 | 2 |
| HR183 | *USP17L22* | M | FCD IIB | Lesionectomy | Left | Temporal | <=5Years | 14/mo | 3 | 3 |
| HR183 | *RBMX* | M | FCD IIB | Lesionectomy | Left | Temporal | <=5Years | 14/mo | 3 | 3 |
| HR184 | *RFX7* | F | FCD IIA | Lobectomy | Left | Temporal | <=5Years | 150/mo | 1 | 1 |
| HR184 | *P2RX5* | F | FCD IIA | Lobectomy | Left | Temporal | <=5Years | 150/mo | 1 | 1 |
| HR184 | *ZACN* | F | FCD IIA | Lobectomy | Left | Temporal | <=5Years | 150/mo | 1 | 1 |
| HR187 | *ZDHHC11B* | M | FCD IIA | Lesionectomy | Left | Temporal | <=5Years | 210/mo | 1 | 1 |
| HR187 | *PCDHB9* | M | FCD IIA | Lesionectomy | Left | Temporal | <=5Years | 210/mo | 1 | 1 |
| HR189 | *PSPC1* | M | HME | Hemispherotomy | Right | Frontal | <=5Years | 100/mo | 1 | 1 |
| HR193 | *SMPDL3B* | M | FCD IIB | Hemispherotomy | Right | Frontal | <=5Years | 750/mo | 1 | 1 |
| HR193 | *DISP1* | M | FCD IIB | Hemispherotomy | Right | Frontal | <=5Years | 750/mo | 1 | 1 |
| HR193 | *OR10A2* | M | FCD IIB | Hemispherotomy | Right | Frontal | <=5Years | 750/mo | 1 | 1 |
| HR193 | *IDO2* | M | FCD IIB | Hemispherotomy | Right | Frontal | <=5Years | 750/mo | 1 | 1 |
| HR193 | *PLEC* | M | FCD IIB | Hemispherotomy | Right | Frontal | <=5Years | 750/mo | 1 | 1 |
| HR196 | *ZNF480* | M | FCD IIB | Lesionectomy | Left | Frontal | <=5Years | Uncontrollable | 2 | 2 |
| HR196 | *SLC29A1* | M | FCD IIB | Lesionectomy | Left | Frontal | <=5Years | Uncontrollable | 2 | 2 |

## Notes

### Competing Interest Statement

The authors have declared no competing interest.

### Author Declarations

This research was approved by the Research Ethics Committee of HCFMRP-USP (Protocol No. 442 6979/2015).

## References

Andrews S. 2010. FastQC: a quality control tool for high throughput sequence data. (No Title).

Arai Y, Edwards V, Becker LE. 1999. A comparison of cell phenotypes in hemimegalencephaly and tuberous sclerosis. Acta Neuropathol 98: 407–413.

Baldassari S, Klingler E, Teijeiro LG, Doladilhe M, Raoux C, Roig-Puiggros S, Bizzotto S, Couturier J, Gilbert A, Sami L et al. 2025. Single-cell genotyping and transcriptomic profiling of mosaic focal cortical dysplasia. Nat Neurosci doi:10.1038/s41593-025-01936-z.

Baldassari S, Ribierre T, Marsan E, Adle-Biassette H, Ferrand-Sorbets S, Bulteau C, Dorison N, Fohlen M, Polivka M, Weckhuysen S. 2019. Dissecting the genetic basis of focal cortical dysplasia: a large cohort study. Acta neuropathologica 138: 885–900.

Baulac S, Ishida S, Marsan E, Miquel C, Biraben A, Nguyen DK, Nordli D, Cossette P, Nguyen S, Lambrecq V. 2015. Familial focal epilepsy with focal cortical dysplasia due to DEPDC 5 mutations. Annals of neurology 77: 675–683.

Benegas G, Albors C, Aw AJ, Ye C, Song YS. 2025. A DNA language model based on multispecies alignment predicts the effects of genome-wide variants. Nature Biotechnology: 1–6.

Berg AT, Zelko FA, Levy SR, Testa FM. 2012. Age at onset of epilepsy, pharmacoresistance, and cognitive outcomes: a prospective cohort study. Neurology 79: 1384–1391.

Béroud C, Collod-Béroud G, Boileau C, Soussi T, Junien C. 2000. UMD (Universal mutation database): a generic software to build and analyze locus-specific databases. Human mutation 15: 86–94.

Blümcke I, Thom M, Aronica E, Armstrong DD, Vinters HV, Palmini A, Jacques TS, Avanzini G, Barkovich AJ, Battaglia G. 2011. The clinicopathologic spectrum of focal cortical dysplasias: A consensus classification proposed by an ad hoc task force of the ILAE Diagnostic Methods Commission 1. Wiley Online Library.

Breuss MW, Yang X, Schlachetzki JC, Antaki D, Lana AJ, Xu X, Chung C, Chai G, Stanley V, Song Q. 2022. Somatic mosaicism reveals clonal distributions of neocortical development. Nature 604: 689–696.

Chessell IP, Hatcher JP, Bountra C, Michel AD, Hughes JP, Green P, Egerton J, Murfin M, Richardson J, Peck WL. 2005. Disruption of the P2X7 purinoceptor gene abolishes chronic inflammatory and neuropathic pain. Pain 114: 386–396.

Chung C, Yang X, Bae T, Vong KI, Mittal S, Donkels C, Phillips HW, Marsh APL, Breuss MW, Ball LL et al. 2023. Comprehensive multi-omic profiling of somatic mutations in malformations of cortical development. Nature Genetics 55: 209–220.

Cingolani P, Platts A, Wang LL, Coon M, Nguyen T, Wang L, Land SJ, Lu X, Ruden DM. 2012. A program for annotating and predicting the effects of single nucleotide polymorphisms, SnpEff: SNPs in the genome of Drosophila melanogaster strain w1118; iso-2; iso-3. fly 6: 80–92.

D’Gama AM, Woodworth MB, Hossain AA, Bizzotto S, Hatem NE, LaCoursiere CM, Najm I, Ying Z, Yang E, Barkovich AJ. 2017. Somatic mutations activating the mTOR pathway in dorsal telencephalic progenitors cause a continuum of cortical dysplasias. Cell reports 21: 3754–3766.

De Angelis L. 1989. Modification of the behavioural effects of amineptine after acute, subacute and chronic administration of phenobarbital in mice. *In Vivo (Athens*, Greece) 3: 331–334.

Elia J, Glessner JT, Wang K, Takahashi N, Shtir CJ, Hadley D, Sleiman PM, Zhang H, Kim CE, Robison R. 2012. Genome-wide copy number variation study associates metabotropic glutamate receptor gene networks with attention deficit hyperactivity disorder. Nature genetics 44: 78–84.

Ewels P, Magnusson M, Lundin S, Käller M. 2016. MultiQC: summarize analysis results for multiple tools and samples in a single report. Bioinformatics 32: 3047–3048.

Frazer J, Notin P, Dias M, Gomez A, Min JK, Brock K, Gal Y, Marks DS. 2021. Disease variant prediction with deep generative models of evolutionary data. Nature 599: 91–95.

Garcia CA, Carvalho SC, Yang X, Ball LL, George RD, James KN, Stanley V, Breuss MW, Thomé U, Santos MV. 2020. mTOR pathway somatic variants and the molecular pathogenesis of hemimegalencephaly. Epilepsia open 5: 97–106.

Garcia CAB, Zubair M, Santos MV, Lee SH, Graham IA, Stanley V, George RD, Gleeson JG, Machado HR, Yang X. 2025. Identification of Novel Mosaic Variants in Focal Epilepsy-Associated Patients’ Brain Lesions. Genes 16: 421.

Garn R, Gul S, Anilkumar A. 2025. Brain Imaging Features in Knobloch Syndrome Type 1 due to COL18A1 Mutation (P2-6.017). In Neurology, Vol 104, p. 1589.

Lippincott Williams & Wilkins Hagerstown, MD. Grajkowska W, Kotulska K, Jurkiewicz E, Matyja E. 2010. Brain lesions in tuberous sclerosis complex. Review. Folia Neuropathol 48: 139–149.

Hunt SE, Moore B, Amode RM, Armean IM, Lemos D, Mushtaq A, Parton A, Schuilenburg H, Szpak M, Thormann A. 2022. Annotating and prioritizing genomic variants using the Ensembl Variant Effect Predictor—A tutorial. Human mutation 43: 986–997.

Jaglin XH, Poirier K, Saillour Y, Buhler E, Tian G, Bahi-Buisson N, Fallet-Bianco C, Phan-Dinh-Tuy F, Kong XP, Bomont P. 2009. Mutations in the β-tubulin gene TUBB2B result in asymmetrical polymicrogyria. Nature genetics 41: 746–752.

Jamuar SS, Lam A-TN, Kircher M, D’Gama AM, Wang J, Barry BJ, Zhang X, Hill RS, Partlow JN, Rozzo A. 2014. Somatic mutations in cerebral cortical malformations. New England Journal of Medicine 371: 733–743.

Jansen LA, Mirzaa GM, Ishak GE, O’Roak BJ, Hiatt JB, Roden WH, Gunter SA, Christian SL, Collins S, Adams C. 2015. PI3K/AKT pathway mutations cause a spectrum of brain malformations from megalencephaly to focal cortical dysplasia. Brain 138: 1613–1628.

Kaswan ZAM, Hurtado M, Chen EY, Steelman AJ, McCusker RH. 2025. Ido1 or Ido2 deficiency in myeloid-derived cells attenuates TMEV-induced ictogenesis. Journal of Neuroimmunology: 578707.

Kircher M, Witten DM, Jain P, O’roak BJ, Cooper GM, Shendure J. 2014. A general framework for estimating the relative pathogenicity of human genetic variants. Nature genetics 46: 310–315.

Li H, Durbin R. 2009. Fast and accurate short read alignment with Burrows–Wheeler transform. bioinformatics 25: 1754–1760.

Lim JS, Kim W-i, Kang H-C, Kim SH, Park AH, Park EK, Cho Y-W, Kim S, Kim HM, Kim JA. 2015. Brain somatic mutations in MTOR cause focal cortical dysplasia type II leading to intractable epilepsy. Nature medicine 21: 395–400.

Liu X, Sun Q, Cao Z, Liu W, Zhang H, Xue Z, Zhao J, Feng Y, Zhao F, Wang J. 2022. Identification of RNA N6-methyladenosine regulation in epilepsy: Significance of the cell death mode, glycometabolism, and drug reactivity. Frontiers in Genetics 13: 1042543.

Mayer C, Hafemeister C, Bandler RC, Machold R, Batista Brito R, Jaglin X, Allaway K, Butler A, Fishell G, Satija R. 2018. Developmental diversification of cortical inhibitory interneurons. Nature 555: 457–462.

McKenna A, Hanna M, Banks E, Sivachenko A, Cibulskis K, Kernytsky A, Garimella K, Altshuler D, Gabriel S, Daly M. 2010. The Genome Analysis Toolkit: a MapReduce framework for analyzing next-generation DNA sequencing data. Genome research 20: 1297–1303.

Mo A, Walsh CA. 2025. Clinical and neuropsychological phenotyping of individuals with somatic variants in neurodevelopmental disorders. Neurology: Genetics 11: e200254.

Møller RS, Weckhuysen S, Chipaux M, Marsan E, Taly V, Bebin EM, Hiatt SM, Prokop JW, Bowling KM, Mei D. 2016. Germline and somatic mutations in the MTOR gene in focal cortical dysplasia and epilepsy. Neurology: Genetics 2: e118.

Najm I, Lal D, Alonso Vanegas M, Cendes F, Lopes-Cendes I, Palmini A, Paglioli E, Sarnat HB, Walsh CA, Wiebe S. 2022. The ILAE consensus classification of focal cortical dysplasia: an update proposed by an ad hoc task force of the ILAE diagnostic methods commission. Epilepsia 63: 1899–1919.

Najm I, Sarnat HB, Blümcke I. 2018. The international consensus classification of Focal Cortical Dysplasia–a critical update 2018. Neuropathology and applied neurobiology 44: 18–31.

Nowakowski TJ, Bhaduri A, Pollen AA, Alvarado B, Mostajo-Radji MA, Di Lullo E, Haeussler M, Sandoval-Espinosa C, Liu SJ, Velmeshev D. 2017. Spatiotemporal gene expression trajectories reveal developmental hierarchies of the human cortex. Science 358: 1318–1323.

Peralta FA, Huidobro-Toro JP. 2016. Zinc as allosteric ion channel modulator: ionotropic receptors as metalloproteins. International Journal of Molecular Sciences 17: 1059.

Poduri A, Evrony GD, Cai X, Elhosary PC, Beroukhim R, Lehtinen MK, Hills LB, Heinzen EL, Hill A, Hill RS. 2012. Somatic activation of AKT3 causes hemispheric developmental brain malformations. Neuron 74: 41–48.

Poduri A, Evrony GD, Cai X, Walsh CA. 2013. Somatic mutation, genomic variation, and neurological disease. Science 341: 1237758.

Program CCS, Abdulla S, Aevermann B, Assis P, Badajoz S, Bell SM, Bezzi E, Cakir B, Chaffer J, Chambers S. 2025. CZ CELLxGENE Discover: a single-cell data platform for scalable exploration, analysis and modeling of aggregated data. Nucleic acids research 53: D886–D900.

Qian X, Coleman K, Jiang S, Kriz AJ, Marciano JH, Luo C, Cai C, Manam MD, Caglayan E, Lai A et al. 2025. Spatial transcriptomics reveals human cortical layer and area specification. Nature 644: 153–163.

Rentzsch P, Witten D, Cooper GM, Shendure J, Kircher M. 2019. CADD: predicting the deleteriousness of variants throughout the human genome. Nucleic acids research 47: D886–D894.

Revel FG, Moreau J-L, Gainetdinov RR, Bradaia A, Sotnikova TD, Mory R, Durkin S, Zbinden KG, Norcross R, Meyer CA. 2011. TAAR1 activation modulates monoaminergic neurotransmission, preventing hyperdopaminergic and hypoglutamatergic activity. Proceedings of the national academy of sciences 108: 8485–8490.

Schreiber V, Dantzer F, Ame J-C, De Murcia G. 2006. Poly (ADP-ribose): novel functions for an old molecule. Nature reviews Molecular cell biology 7: 517–528.

Steriade C, Titulaer MJ, Vezzani A, Sander JW, Thijs RD. 2021. The association between systemic autoimmune disorders and epilepsy and its clinical implications. Brain 144: 372–390.

Tate J. 2010. COSMIC: the catalogue of somatic mutations in cancer. Nucleic Acids Res 38: 164.

Vahidnezhad H, Youssefian L, Harvey N, Tavasoli AR, Saeidian AH, Sotoudeh S, Varghaei A, Mahmoudi H, Mansouri P, Mozafari N. 2022. Mutation update: The spectra of PLEC sequence variants and related plectinopathies. Human mutation 43: 1706–1731.

Zubair M, Yang X. 2025. Dipping Into the Phenotypic Implications of Mosaic Variants. Vol 11, p. e200256. Wolters Kluwer Baltimore.

